# Feature Selection with Quantum Annealing for Biomedical Machine Learning Applications

**DOI:** 10.64898/2026.07.02.26357174

**Authors:** Sarah N. Dudgeon, Seung Joo Lee, Thomas JS Durant, Brent Nelson, H. Patrick Young, Lucila Ohno-Machado, R. Andrew Taylor, Wade L. Schulz

**Author notes:** Corresponding Author: 55 Park St, New Haven, CT 06510. These authors contributed equally to this work (Sarah N. Dudgeon, Seung Joo Lee).

## Abstract

Feature selection is a commonly used method in biomedical artificial intelligence and machine learning to identify a subset of high-quality variables that can be used to train downstream predictive models. It has been suggested that quantum feature selection (QFS), which takes advantage of the properties of quantum computers, may better identify variables that are correlated with the outcome while simultaneously reducing redundancy between selected variables. However, there are a limited number of studies evaluating their performance, particularly in real-world data sets. Here, we assess the performance of two QFS methods compared to random forest (RF) feature selection based on feature stability and the performance of a downstream classification algorithm when used to predict urinary tract infections in the emergency department from 211 original features extracted from the electronic health record. We found that a quantum binary quadratic model (BQM) and constrained quadratic model (CQM) had similar performance to RF feature selection (median F1 score of 0.60, 0.61, and 0.61 respectively) when 10 features were selected for an XGBoost classification model. The BQM and RF also had similar feature stability (0.91 and 0.94, respectively) while the CQM had lower stability (0.72). These findings show that QFS can be used with large, clinical data sets to identify features with high stability and predictive performance. As the capacity and quality of quantum computers continue to increase, these methods may offer additional benefits to classical feature selection methods.

## Introduction

The use of artificial intelligence and machine learning (AI/ML) in biomedicine continues to grow rapidly, with applications from basic science to clinical care.^1,2^ To realize the potential of AI/ML in clinical predictive models, algorithms with high predictive performance are needed, similar to what is seen with in vitro diagnostics. While there is a common assumption that adding more features increases model performance, numerous studies have shown that prioritizing the quality rather than the sheer quantity of features often leads to more robust and generalizable results.^3–5^ This principle is particularly relevant when using data from the electronic health record and similar systems, where a large number of features can often be extracted.^4^ This can result in increased noise within the data, increased computational burden, and more complex models that are prone to overfitting and reduced model interpretability.^6,7^ Feature selection methods can help mitigate these issues by identifying a subset of features most strongly associated with the outcome, reducing noise and redundancy and, in some cases, improving the performance of subsequent predictive models.^5,8–10^

Several methods can be used to reduce the number of features used to train machine learning models.^7,11,12^ Filter methods, such as the t-test and mutual information, select features prior to their use in a downstream machine learning algorithm or predictive task. Wrapper methods use a machine learning algorithm, such as a random forest, to identify important features as part of the predictive task but can also be separated from the downstream machine learning method used. Lastly, embedded methods choose features during the training of a machine learning model and are specific to the method being used. However, while many of these methods identify input features that are statistically correlated with the outcome, not all decrease redundant information between features, which can lead to bias in downstream algorithms.^13^

Quantum computers, which rely on quantum bits (qubits) and use the properties of superposition and entanglement, offer potential benefits for feature selection compared to classical algorithms.^13–15^ Unlike classical algorithms, quantum feature selection (QFS) methods simultaneously identify features with strong statistical associations to the outcome while minimizing redundancy among chosen variables.^13,15^ This balance and simultaneous computation may decrease the bias and improve the generalizability of downstream predictive models that use quantum-selected features. While many quantum computing applications have only been evaluated in proof-of-concept studies due to the limited size of current quantum computers, the design of QFS algorithms allows for their practical use in generally available quantum annealing systems.^14,15^ For example, the D*Wave Advantage platform that is commercially available contains over 5,000 qubits. As a result, QFS presents a viable, near-term opportunity to leverage emerging quantum technologies for practical machine learning applications. Quantum feature selection algorithms can be implemented in this platform with either a binary quadratic model (BQM), which encodes a quantum unconstrained binary optimization (QUBO) problem, or a constrained quadratic model (CQM), which encodes a quadratic objective.^15,16^

In this work, we compare classical and quantum feature selection methods using a large, publicly available real-world dataset previously used to predict urinary tract infections in an emergency department (ED) population as a representative clinical example.^17^ We assess both the predictive performance and the stability of selected features under different conditions, aiming to elucidate the current practical value of QFS and its potential advantages over conventional approaches.

## Methods

### Data Set and Data Preprocessing

Data were obtained from a previously released, de-identified dataset used for the prediction of UTIs from an adult ED population.^17^ Briefly, these data were retrospectively collected from four care sites within a single healthcare system for ED visits between March 2013 and May 2016. Patients included adults (>= 18 years) with a urine culture during an ED visit where symptoms associated with UTI were present. For each visit, extracted data included demographics, chief complaint, vitals, medication list, medical history, history and physical exam findings, and selected laboratory tests available from the time of ED entry to either discharge or admission, resulting in 211 features and 80,387 total records. Data were processed using Python (version 3.9) and the Pandas library (version 2.2.2). Categorical variables were one-hot encoded using Pandas. For both classical and quantum feature selection, we assessed the impact of scaling numerical features using no scaling, scaling from –1 to 1, and scaling from 0 to 1. A holdout dataset consisting of 20% of records (n=16,078) was excluded from the feature selection step after pre-processing to prevent data leakage when evaluating the downstream predictive model performance (Figure 1A) using a stratified shuffle split. For each feature selection method described below, we identified the top 10, 20, 30, or 50 features and assessed performance in the downstream classification model.

**Figure 1.**
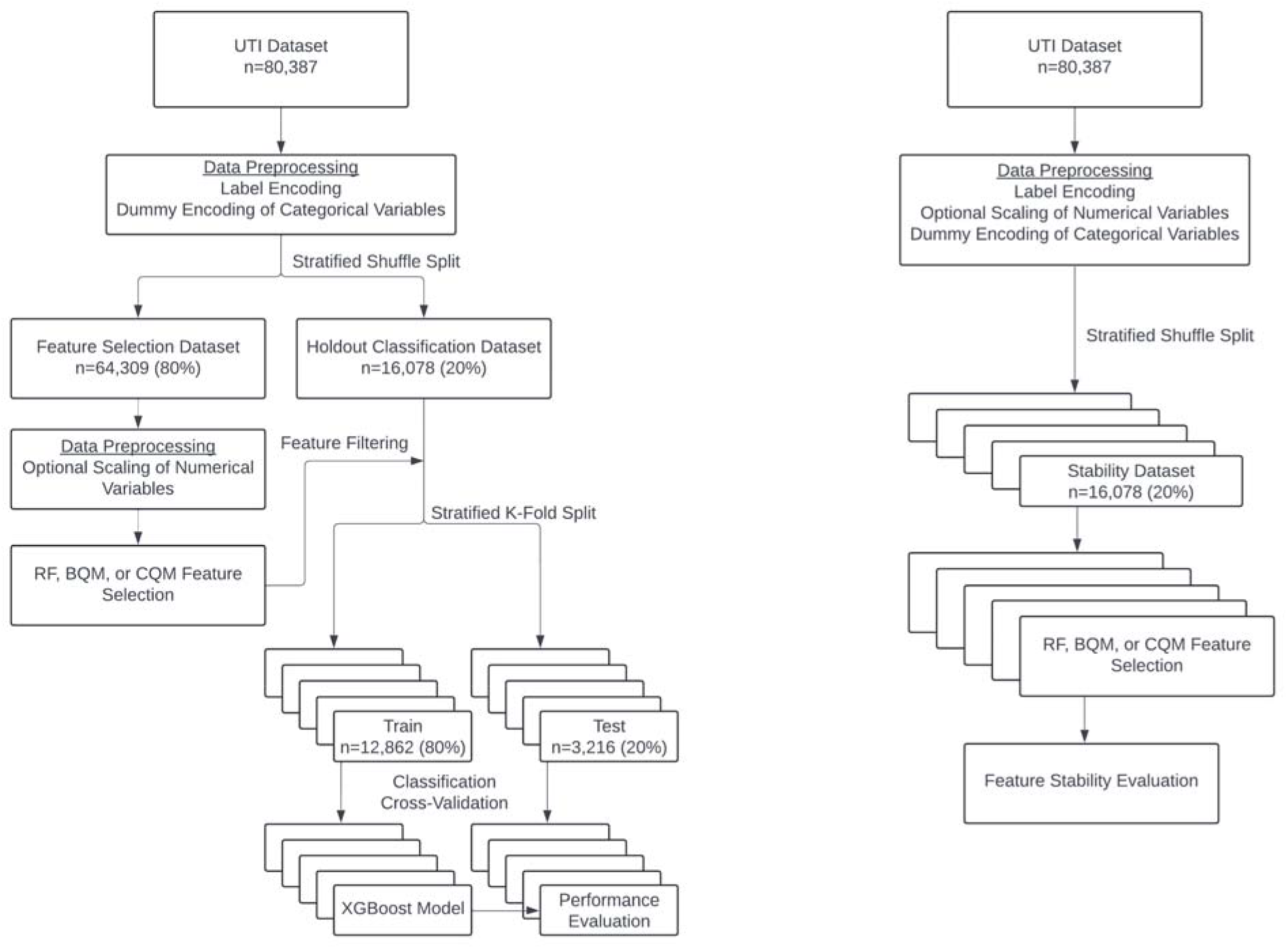

### Classical Feature Selection

We evaluated random forest as a prototypic classical feature selection method based on variable importance. This model was implemented in Python using the Scikit-Learn library (version 1.5.1). We assessed the impact of hyperparameter optimization for the number of estimators and the minimum samples per split during training of the feature selection model (Supplemental Table 1).

### Quantum Feature Selection

In this study, we employed a quantum annealing approach for feature selection using a D-Wave Quantum Processing Unit (QPU). Following initial data preprocessing, we calculated the mutual information (MI) values between each feature and the target variable to assess feature relevance, followed by the conditional mutual information (CMI) between feature pairs to evaluate redundancy (Supplemental Figure 1). These values were then embedded into a feature interaction graph that was mapped into a Quadratic Unconstrained Binary Optimization (QUBO) problem formulation and subsequently mapped to the QPU’s hardware architecture. The QUBO was implemented based on previously published Binary Quadratic Model (BQM) and Constrained Quadratic Model (CQM) algorithms.^18,19^

The BQM model was implemented using the D*Wave Ocean library (version 7.0.0) and the CQM model was implemented with the D*Wave SciKit-Learn plugin (version 0.1.0). The QFS methods were executed on the D*Wave Leap Advantage System (version 5.4) which has over 5,000 qubits with 15-way connectivity.^16^ Because of the limited size and qubit connectivity, large feature sets like those in our benchmarking dataset could not be directly modeled using the BQM formulation. The number of input features was therefore reduced based on mutual information (MI) and conditional mutual information (CMI) scores, and a subset of the most strongly correlated features were provided to the quantum algorithm. For the BQM model, we optimized BQM strength, the number of QPU reads, the number of Kerberos sampler iterations, and the number of initial features provided to the QFS algorithm (Supplemental Table 1). For the CQM model, we evaluated the impact of varying the alpha bias coefficient (Supplemental Table 1).

### Performance Evaluation

The performance of each feature selection method was determined by assessing the ability of a gradient-boosted descent classifier to accurately predict positive urine cultures. This model was implemented in Python using the XGBoost library (version 2.1.0). For classification model training and evaluation, a 5-fold cross-validation using a stratified k-fold split (k=5) from the holdout dataset was performed (Figure 1A). Performance metrics (F1 score, precision, specificity, accuracy, and balanced accuracy) were calculated using the Scikit-Learn library for each combination of configuration parameters. For the BQM method, it is not guaranteed that the exact number of features specified will be returned; therefore, results that returned a different number of features were excluded from the performance evaluation.

For the top performing RF, BQM, and CQM models for 20 features, feature stability was calculated. For this evaluation, a 5-fold stratified shuffle split was performed and the top features identified for each split (Figure 1B). Feature stability was calculated using the Python stability library developed by Nogueira et al.^6^ Experiment results were logged using the Azure Machine Learning/mlflow library (version 1.56.0).

### Data and Code Availability

Data are available from the original publication.^17^ All codes were written and independently reviewed by separate authors and are available at https://github.com/acomphealth/quantum-feature-selection.

## Results

### Quantum Feature Selection Baseline Comparison

We first assessed the downstream classification performance obtained using features selected by each method and the set of configuration parameters that produced the highest median F1 score in the holdout dataset (Figure 1A). For all feature selection methods, the performance of the classification model improved as the number of selected features increased (Figure 2). Overall, the performance of the classical (RF) and quantum (BQM, CQM) feature selection methods was similar across feature subset sizes. Using the top 10 selected features, the RF achieved a median F1 of 0.61, the quantum BQM achieved 0.60, and the quantum CQM achieved 0.61 for downstream classification performance. When the top 50 selected features were used, the RF achieved a median F1 of 0.67, the quantum BQM achieved 0.66, and the quantum CQM achieved 0.65.

**Figure 2.**
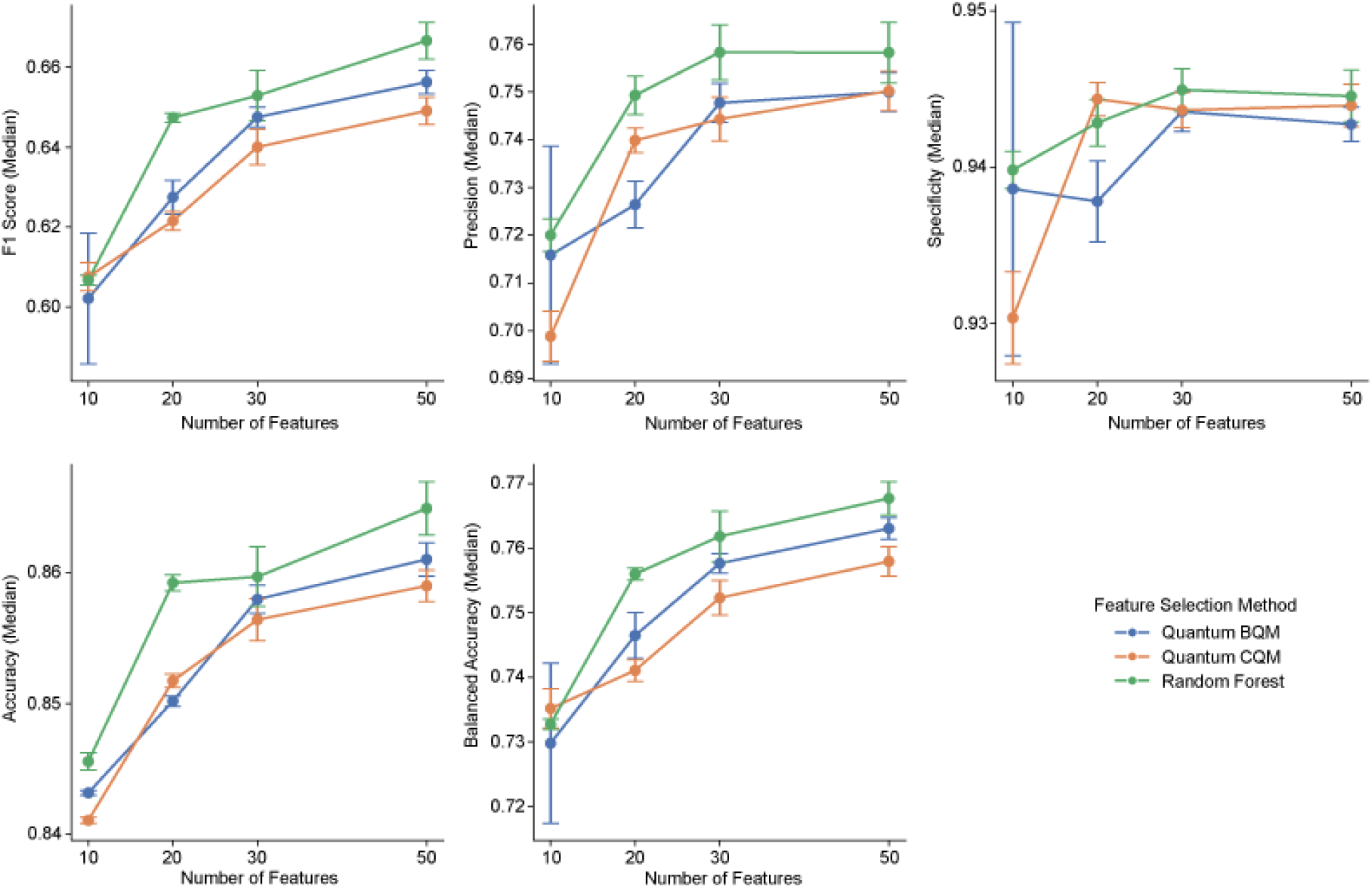

### Optimization of Quantum Feature Selection Models

The selection of configuration parameters for data preprocessing and model hyperparameters influenced the performance of the feature selection methods, particularly when selecting a smaller number of features. The CQM method showed the greatest variability in downstream classification performance, with median F1 scores ranging from 0.43 to 0.62 when using the top 10 features and from 0.59 to 0.65 with 50 features (Figure 3A). The other models showed less variation across trials, with the BQM model having a range of 0.55-0.60 for 10 features and 0.65-0.66 with 50 features (Figure 3B). The RF model had a median F1 range of 0.58-0.61 with 10 features and 0.66-0.67 with the top 50 features (Figure 3C).

**Figure 3.**
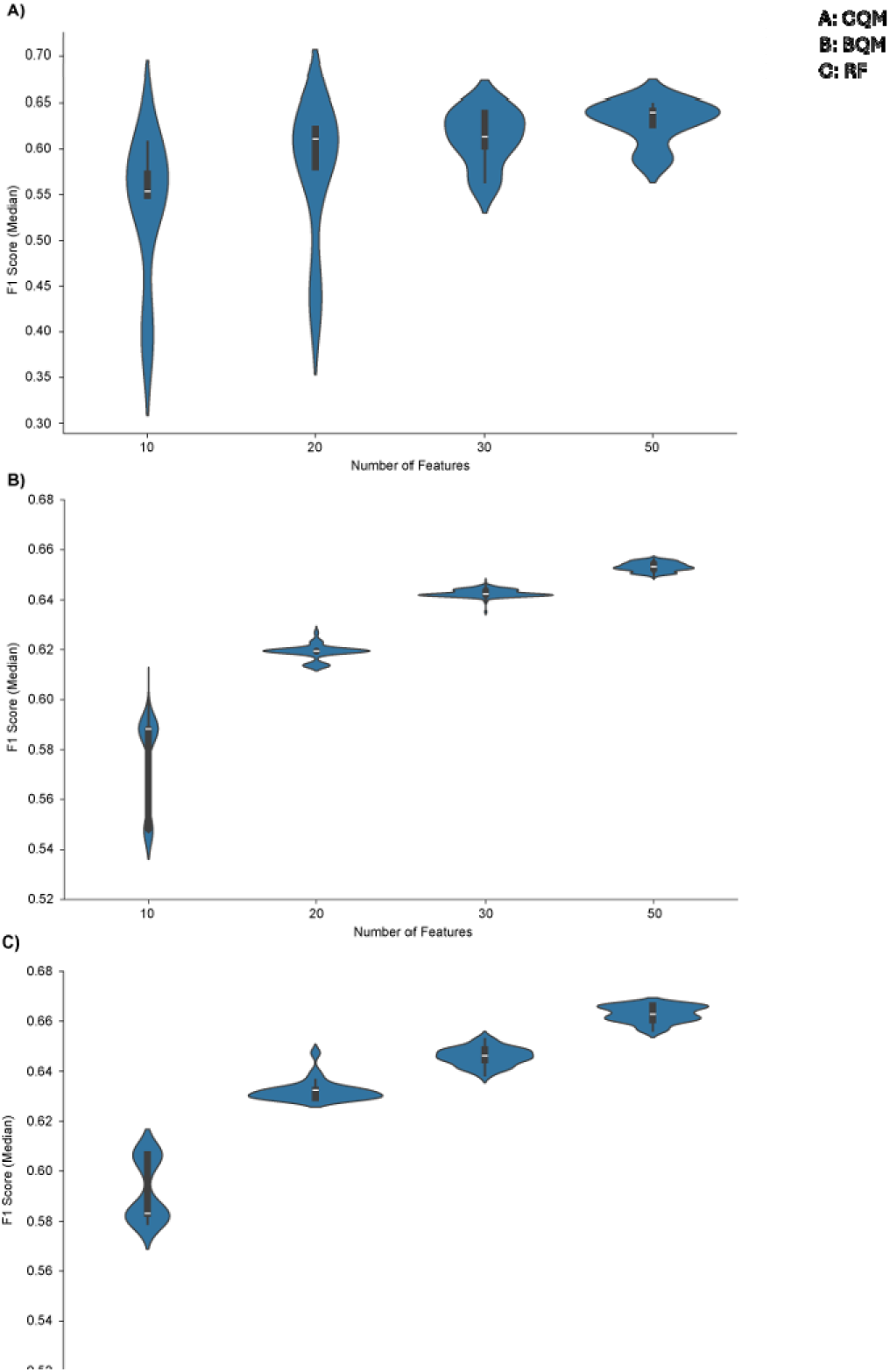

We also evaluated the impact of individual configuration parameters on classification performance. The choice of data scaler had minimal impact on the performance for either the RF or BQM models (Table 1). Based on this finding, only a single scaler was used to evaluate the CQM model to conserve quantum computing resources. For the CQM model, the alpha parameter was the most important configuration parameter (Table 1).

**Table 1.**
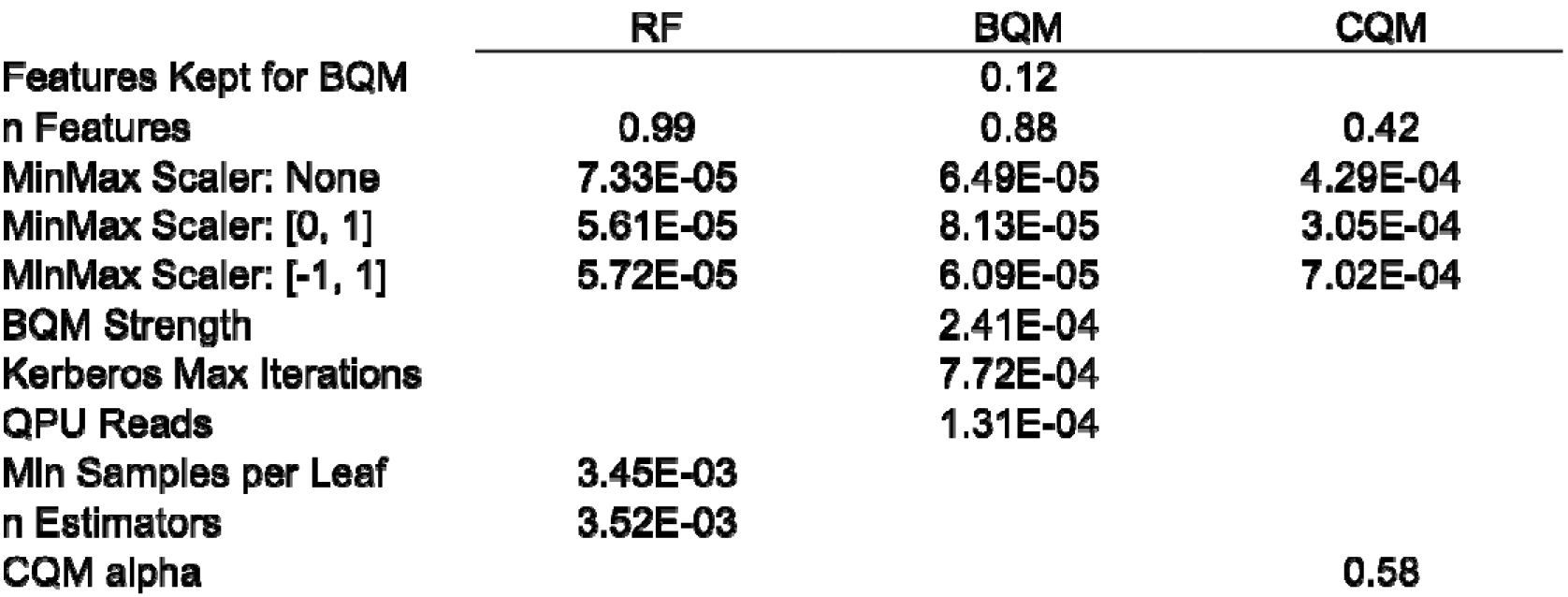

The BQM method has two model-specific configuration parameters, the number of features provided to the model and the penalty strength to enforce the feature subset size. As shown in Table 1, the number of features provided from the overall feature set to the BQM model was an important configuration parameter that influenced downstream performance. Since the BQM model is limited to variables that have a known statistical correlation with the label, we next wanted to verify that the BQM method identified features with better performance than the mutual information method alone. As shown in Figure 4A, performance improved when more than the minimum number of features was provided, indicating that the BQM method selected more informative, better performing features than the mutual information method alone.

**Figure 4.**
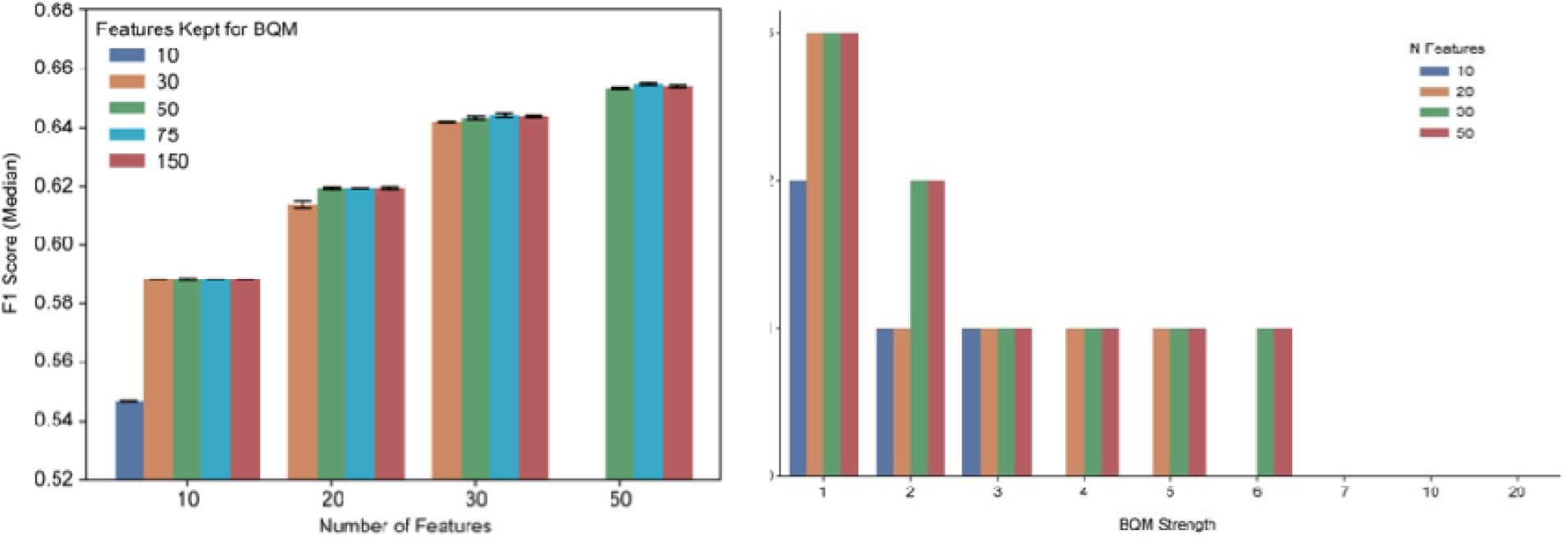

The second BQM-specific configuration parameter is the BQM strength, which functions as a penalty score to ensure that the model returns the exact number of features requested. Figure 4B shows the impact of increasing penalty scores on the number of features returned by the model compared to the number of features requested. As the number of requested features increases, higher penalty strengths were required to ensure that only the top n features are returned.

### Stability of Classical and Quantum Feature Selection Methods

Feature stability was evaluated to assess the consistency of features identified by each feature selection method. We found that the RF and BQM models both had high stability in a 5-fold stratified shuffle split of the data when selecting 20 features (Table 2). The CQM model showed lower feature stability than both RF and BQM.

**Table 2.**

|  | **Stability** | **Variance** |
| --- | --- | --- |
| **RF** | **0.94** | **4.69E-05** |
| **BQM** | **0.91** | **2.52E-04** |
| **CQM** | **0.72** | **3.41E-05** |

## Discussion

In this study, we found that QFS methods using commercially available quantum annealing systems can achieve similar performance to a classical Random Forest based approach. In addition, the BQM method demonstrated feature selection stability similar to RF. As with other machine learning methods, configuration parameters and hyperparameters should be evaluated to assess and potentially improve the overall performance of feature selection methods. While not all parameters evaluated here demonstrated large variations in model performance, these findings may be dataset dependent. Our findings suggest that quantum annealing based feature selection can produce robust and clinically meaningful feature subsets comparable to those identified by classical approaches despite current hardware limitations when appropriate model parameter configurations are used.

Feature selection is inherently a combinatorial optimization problem, as the number of possible feature subsets increases exponentially with the number of candidate variable^20^. Classical approaches such as RF based feature importance largely rely on heuristic or greedy search strategies to identify informative features and may become trapped in local optima and fail to reach global optimum when the search space contains complex interactions among variables^21^. Quantum computing shows promise for addressing complex computational challenges including feature selection in healthcare. While gate-based quantum systems are limited in available qubits, quantum annealers can currently reach over 5,000 qubits, making them better suited for complex real-world optimization tasks. Quantum annealers take advantage of quantum tunneling that avails exploring through rugged optimization landscapes and escaping local minima^22^. Assessing the performance of quantum approaches relative to established classical feature selection methods such as RF can therefore help determine the promise and practical advantages of these emerging approaches.

Other preliminary work has shown that QFS may outperform classical methods.^13^ However, this prior study did not compare the QFS methods to any classical machine learning approaches for feature selection, which may perform better with larger data sets^23^, and several of the datasets evaluated had substantial class imbalance. Other studies have demonstrated that informative features can be identified using gate-based quantum algorithms, but reported lower performance compared to classical methods, likely influenced by restrictions on data set size and feature dimensionality imposed by limitations in currently available quantum hardware capacity.^24^ Our findings show that QFS methods achieved high-quality feature selection on a large, real-world data set derived from the electronic health record consisting of over 80,000 records and 211 original features with comparable performance to the RF-based importances. While we also needed to reduce the number of QFS input features using classical statistical methods for the BQM method, our results demonstrated that the quantum model improved the quality of features selected beyond the classical mutual information and conditional information values alone based on the increase in classification F1 score when more features were provided to the quantum BQM method (Figure 4A). These findings support other studies using quantum annealing systems for feature selection from medical imaging data, validating that the quantum computer may contribute additional value beyond the classical filtering step.^25^

While the CQM achieved similar downstream classification performance compared to the RF model or BQM, it showed lower feature stability as well as more variable performance. This may be related to the increased complexity of the CQM solver. CQM is designed to accommodate more complex optimization problems by supporting additional, hard constraints and feature types with a larger problem space^26^. While this increased modeling flexibility allowing for all features to be evaluated directly in the quantum system in our experiments, it also created a more complex energy landscape with multiple solutions of similar objective values, reducing consistency in the solutions identified across data splits. Another possible explanation for its lower stability is that hard constraints in the CQM are typically incorporated into the objective function as penalties with predefined weights. As a result, soft constraint violations may occur in some cases when the solver prioritizes reaching a lower energy state (objective optimization) over strictly satisfying all constraints. Consistent with this, the alpha parameter, which controls the relative importance of the constraint (number of features) in the CQM formulation, showed the largest influence among the CQM configuration parameters evaluated in our study (Table 1). In contrast, the number of selected features had the largest coefficients in RF and BQM models. As a result, the CQM may identify different but similarly performing feature subsets across data splits, resulting in lower observed feature stability.

The CQM also consumed more quantum resources using the hybrid solver (approximately 70 seconds per experiment). The BQM was computationally more efficient, consuming minimal quantum computing time for each trial (<180 milliseconds per experiment). These findings highlight the practical tradeoff between optimization flexibility, reproducibility, and computational efficiency in applying a quantum feature selection method under current hardware constraints. Based on literature and our findings, the BQM appears to be more reliable and suitable for smaller optimization problems, whereas the CQM can handle larger and more complex problems but require careful formulation and appropriate scaling of configuration parameters such as constraint penalty weights and bias.

Despite comparable performance and feature stability, neither quantum model consistently outperformed the classical model in this study. These results suggest that quantum annealing based feature selection shows promise as an alternative optimization framework, though further investigations may be required to confirm the practical advantages of these approaches using currently available quantum hardware. This aligns with the broader challenge of implementing machine learning models in healthcare, where trust, interpretability, and computational efficiency often constrain model design.^2,7^ Many clinical prediction models rely on a small number of features to support scoring algorithms, even when additional data might improve performance. Notably, Taylor et al., who released the dataset used in this work, demonstrated that a set of 10 expert-selected features could match the performance of a model trained on all 211 original variables.^17^ In this study, we found that automated feature selection methods, both classical and quantum, can be used to train accurate predictive models. Specifically, our findings demonstrate that quantum annealing based feature selection can achieve performance comparable to established classical methods on real-world datasets, highlighting its potential as a scalable optimization framework as quantum hardware continues to evolve.

## Conclusion

Quantum feature selection methods were able to achieve equivalent performance to classical RF importance to predict positive urine cultures using a limited subset of clinical data. While we did not find an overall improvement in classification performance using QFS for this dataset, as the field of QML continues to evolve and quantum computers grow in capacity and sophistication, these approaches may offer advantages to their classical counterparts. Quantum feature selection methods offer an immediately available methodology that can be used with large data sets, but further research will be needed to identify which types of data may benefit most from the applications of these algorithms.

## Data Availability

Data are available from the original publication.All codes were written and independently reviewed by separate authors and are available at https://github.com/acomphealth/quantum-feature-selection.

---

**Supplemental Figure 1.**
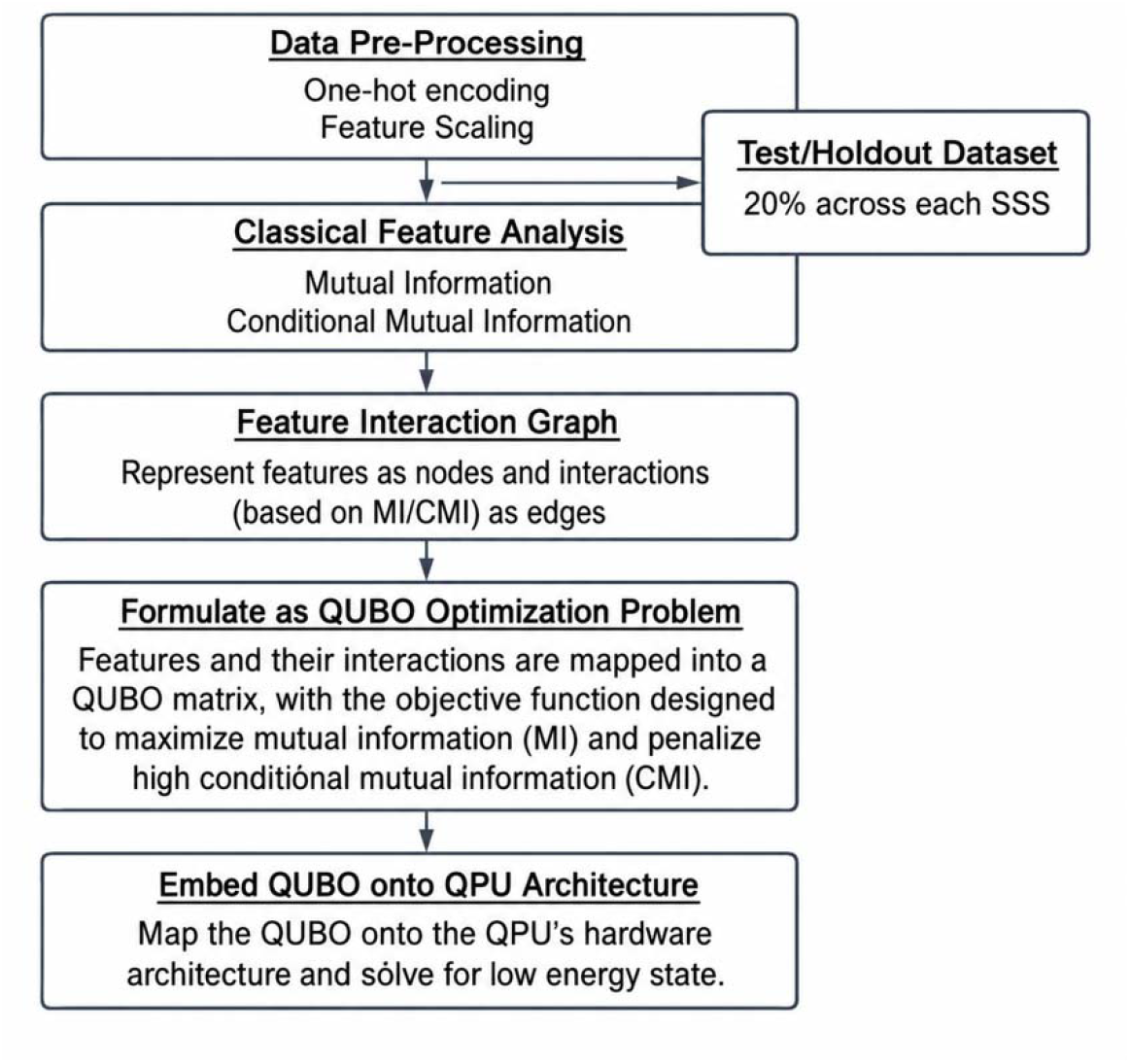

**Supplemental Table 1.**
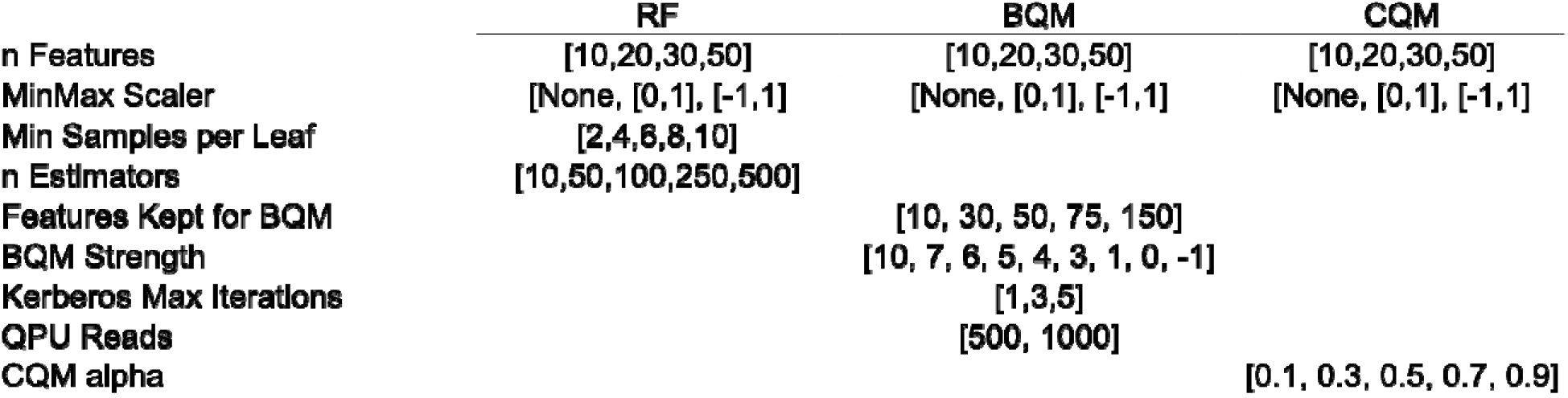

